# Using collaboration networks to identify authorship bias in meta-analyses

**DOI:** 10.1101/19001305

**Authors:** Thiago C. Moulin, Olavo B. Amaral

## Abstract

Meta-analytic methods are powerful resources to summarize the existing evidence concerning a given research question, and are widely used in many academic fields. However, meta-analyses can be vulnerable to various sources of bias, which should be considered to avoid inaccuracies. Many of these sources can be related to study authorship, as both methodological choices and researcher bias may lead to deviations in results between different research groups. In this work, we describe a method to objectively attribute study authorship within a given meta-analysis to different research groups by using graph cluster analysis of collaboration networks. We then provide empirical examples of how the research group of origin can impact effect size in distinct types of meta-analyses, demonstrating how non-independence between within-group results can bias effect size estimates if uncorrected. Finally, we show that multilevel random-effects models using research group as a level of analysis can be a simple tool for correcting biases related to study authorship.

## Introduction

The scientific process is prone to several types of bias that can undermine the reliability of the research literature^1^. The origins and consequences of this problem have been extensively described; however, attempts at solutions have so far been insufficient, as recent analyses of the literature indicate that issues such as publication^2,3^ and sponsorship^4^ biases are still widespread. Moreover, intrinsic aspects of the current publication, peer-review and reward systems have been shown to lead to bias towards overly positive and inflated results^5–7^.

As a consequence of bias in the original studies, summarizations and meta-analysis of the existing literature can lead to misleading outcomes^8,9^. Moreover, the meta-analytic process itself can be biased by the selective inclusion of studies^10^. On the other hand, meta-analyses can also be used to detect and quantify sources of bias. A number of methods have been created for this purpose, focusing mainly on publication and reporting biases^11,12^, as well as study quality assessment^13^. However, other sources of bias have received less attention, and new approaches are needed for their systematic study.

A possible source of bias in meta-analyses is the non-independence between study results, which violates the assumptions usually required by statistical models used for data synthesis. When groups of non-independent results are easily identifiable (e.g. outcomes from the same experiment or experiments within the same article), these can be accounted for by diverse methods^14^ When analyzing articles containing several experiments or cohorts, using multilevel models is a way to consider the dependencies within articles^15^. If different outcomes from the same subjects within an experiment are included, leading to non-independence between sampling errors as well, multivariate meta-analyses can be applied^16^. Nevertheless, other sources of non-independence can be harder to detect or approach objectively.

The research group of origin of a study is an obvious source of non-independence between results. Certain authors or groups might be more prone to find certain outcomes, either due to methodological factors (i.e. use of particular protocols, methods or populations) or to biases in performing, analyzing or reporting experiments^17^. As different research groups will not contribute equally to a meta-analysis, this phenomenon, which we will refer to as authorship bias, can potentially distort meta-analytical results. Nevertheless, objective detection of authorship bias is hampered by the lack of a clear definition of what constitutes a research group. As academic mobility is high, collaboration is frequent and authorship criteria are flexible, it is unlikely that two sets of studies from a group will have exactly the same set of authors. At the same time, it is not clear at what point differences between author lists become large enough to attribute studies to different groups.

In this work, we describe a straightforward method to define research groups based on collaboration graphs, which can be used to assess and quantify authorship bias in a meta-analysis. To demonstrate its usefulness, we apply this procedure in different meta-analyses to show that results coming from the same research group can impact results in various ways, leading to potential misinterpretations of the data. We then demonstrate how the use of multilevel random-effects models based on author networks can correct effect size estimation in these cases. The use of these tools might not only increase precision in data synthesis, but also provide a window to study the impact of authorship on results in different fields of research.

## Methods

### Selection of meta-analyses examples and data extraction

As shown in the study outline presented in **Fig. 1**, we extracted data from four meta-analyses to test our method for research group definition and evaluation of authorship bias. We chose meta-analyses from different fields of biomedical science (e.g. clinical trials, cross-sectional studies in humans, experimental animal studies) with open or available raw data as examples, but did not use systematic sampling in this process. The studies are referred to in the text by their article reference, although the specific meta-analyses analyzed were usually one of many included in the original studies (**Table 1**). The first one, from Chen et al.^18^, describes the effects of eye-movement desensitization and reprocessing therapy on the symptoms of posttraumatic stress disorder. Mathie et al.^19^ performed a meta-analysis on double-blind, placebo-controlled trials of homeopathic treatment. Kredlow et al.^20^ studied the post-retrieval extinction effects on fear memories of rodent models. Finally, Munkholm et al.^21^ estimated levels of BDNF in bipolar disorder patients (irrespective of affective state).

**Table 1.** Features of included meta-analyses. Table shows the reference for each meta-analysis (including the corresponding figure in the original article) and the following features: number of articles, number of results, indicators of heterogeneity (Q-test p values and I^2^ values), and small-study effects (Egger’s regression z and p values, as the number of missing studies in trim-and-fill analysis), calculated by the R metafor package.

| Reference | Chen et al.<br>(Fig. 2)<sup>18</sup> | Mathie et al.<br>(Fig. 2)<sup>19</sup> | Kredlow et al.<br>(Fig. 2)<sup>20</sup> | Munkholm et al.<br>(Fig. 1S)<sup>21</sup> |
| --- | --- | --- | --- | --- |
| # articles | 22 | 51 | 10 | 32 |
| # results | 22 | 54 | 34 | 34 |
| Q statistic | 65.14 | 266.43 | 170.08 | 351.71 |
| Q-test p-value | <0.001 | <0.001 | <0.0001 | <0.0001 |
| I<sup>2</sup> (%) | 68.04 [44.3,85.1] | 62.74 [45.7,78.3] | 81.90 [72.9,90.6] | 93.61 [90.2,96.5] |
| Egger's test (z) | -0.2457 | -3.0815 | 1.1882 | -2.9591 |
| Egger's p-value | 0.8059 | 0.0018 | 0.2348 | 0.0011 |
| Trim-and-fill<br>missing studies | 0 | 17 | 1 | 0 |

**Figure 1.**
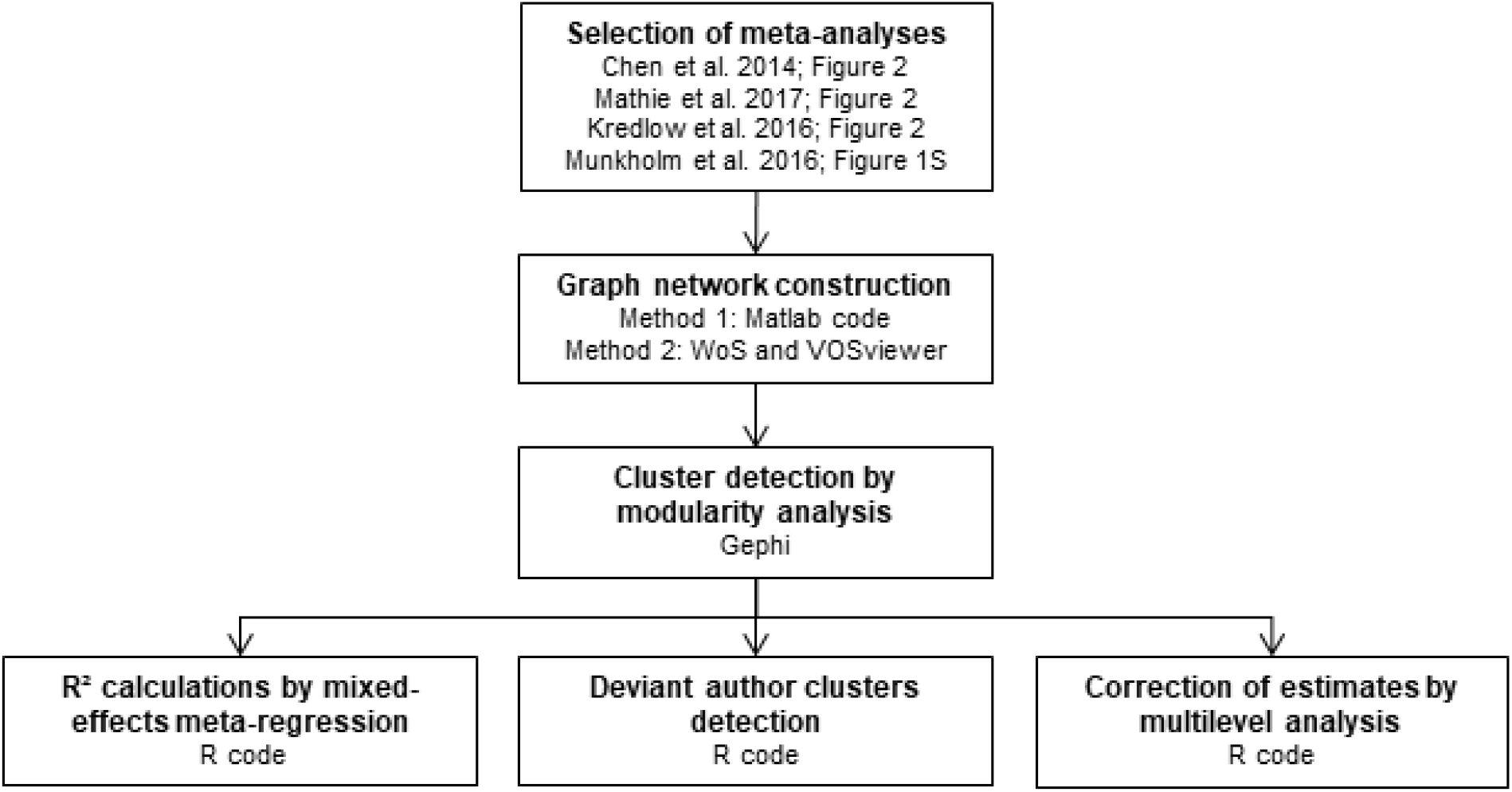
Study flow diagram. After study selection, we used modularity algorithms from graph networks to define author clusters within each meta-analysis. We then adopted different approaches to measure authorship bias: i. Evaluating the influence of author cluster in meta-analysis heterogeneity by R^2^ estimation through mixed-effects meta-regression; ii. Detecting clusters with results diverging from the remaining studies by a Wald-type test between estimates from random-effects models of the cluster and of the aggregate of the remaining ones; iii. Using multilevel analysis to correct meta-analytic estimates. Software tools used are shown for each individual step. CMA (Comprehensive Meta-Analysis v3); Gephi (version 0.9.2); Matlab (MathWorks MATLAB 2017b); R (R-3.5.2 on RStudio 1.1.463); VOSviewer (version 1.6.11); WoS (Web of Science database).

We obtained the effect size, sample size and standard error for each study from figures in the articles (Fig. 2 in Chen et al., 2014, Kredlow et al., 2016 and Mathie et al. 2017; Fig. 1S in Munkholm et al., 2016) except for Kredlow et al., 2016, in which standard error data was obtained by contact with the first author. The data extracted from the figures was fed into the Comprehensive Meta-Analysis version 3.3 (CMA, Biostat Inc.), which converted the data, when necessary, to Hedges’ g estimates and computed sampling variances. From the reference sections, we obtained the PubMed ID (or DOI, when PubMed ID was not available) of the original studies included in the meta-analyses, which we used to generate author networks for each of them. We used the R metafor package to obtain estimates of between-study variance (I^2^ for quantification and Q-test for hypothesis testing), as well as indicators of small-study effects suggestive of publication bias (Egger’s regression and trim-an-fill-analysis, with funnel plots presented in **Supp. Fig. 1**). The choice of trim-and-fill was made following the analyses used in the original articles, but currently there are other recommended options, as selection model approaches. ^22,23^The original features of these meta-analyses can be found in **Supp. Table 1**. Note that the effect sizes in the original studies may diverge from our calculations due to the use of different estimators, as we chose to use a uniform approach for all meta-analyses rather than following the original models.

**Figure 2.**
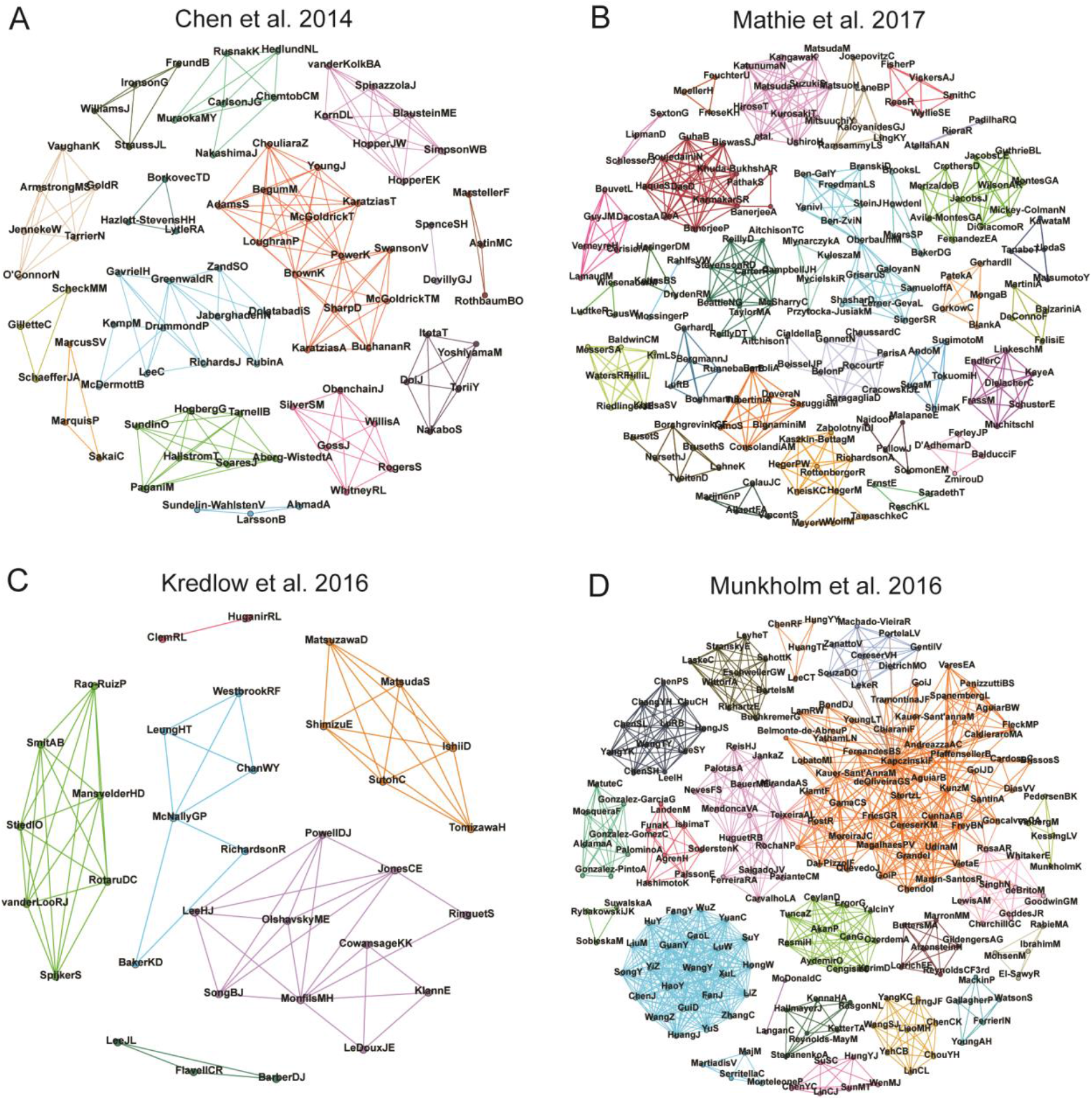
Author networks. The complete network of authors for each meta-analysis used in the study is shown, with nodes representing authors and edges representing collaborations between them within the meta-analysis. Edge weights, defined by the number of coauthored studies between an author pair, are not shown in the figure, but are considered when performing modularity analysis. Clusters emerging from this process are shown in different arbitrary colors. **(A)** Chen et al., 2014 (83 nodes, 195 edges, 16 clusters); **(B)** Mathie et al., 2017 (183 nodes, 467 edges, 40 clusters); **(C)** Kredlow et al., 2016 (34 nodes, 72 edges, 6 clusters); **(D)** Munkholm et al., 2016 (202 nodes, 1010 edges, 21 clusters).

### Construction of author networks

We developed two methods for the construction of the graph networks describing connections between authors (**Fig. 2**): (a) a MATLAB code, available as supplementary material, that uses the PubMed ID or DOI of the original studies and accesses PubMed to search for the authors of each study, connecting those with common publications within the meta-analysis; and (b) manual search of articles in the Web of Science database and data processing with VOSviewer software^24^. Both methods are described in detail hereafter.

### MATLAB code

Network creation using the MATLAB code uses a list of each result in the meta-analysis and either the respective PubMed ID or DOI of its study of origin as input. This information is used to search PubMed for the author list of each article. A list of authors and related study identifiers is then created by the code. If there is no match for a specific search, the DOI number will be listed as an author by itself, which will ultimately become a cluster with no connections. The code uses this output to generate a relationship adjacency matrix of the searches, weighing every connection between authors by the number of co-authored results within the meta-analysis. Both the list of authors and the matrix are saved as CSV files. All routines are available as supplementary material with running examples and brief instructions.

### VOSviewer software

In order to increase the accessibility of our method, we also explored other software resources for alternative ways to build authorship networks. For this, we manually searched the Web of Science database using the PubMed ID (or DOI, when PubMed ID was not available) of all articles in the meta-analysis (a search string example for Munkholm et al. is provided as supplementary data). The retrieved results were saved as a non-formatted text file for VOSviewer handling. In the software, we chose the option of creating a map based on bibliographic data to generate a co-authorship network. Software options were set to (a) full counting (so that each co-authorship would weigh equally), (b) not ignoring documents with large number of authors, and (c) reducing first names to initials. We did not use any minimum threshold for number of publications or citations per author. The output was saved as a GML file.

An advantage of this method compared to the MATLAB code is that it allows the use of other databases besides PubMed, such as Web of Science and Scopus. Moreover, it may be more user-friendly to some researchers. Despite minor differences, both methods achieved a similar number of clusters in our example search (**Supp. Fig. 2**). However, in VOSviewer there is no automatic handling of search errors (i.e.: not finding a DOI number) and the methods to weigh connections between authors are different (i.e.: edges are weighed by the number of shared articles in VOSviewer, while our MATLAB code weighs them by shared results within the meta-analysis; these methods may diverge when there are multiple results within articles), which can cause some changes in clustering. Thus, we decided to use the MATLAB-generated networks for further analyses.

### Lifetime PubMed connections

When exploring ways to consider author networks, we also attempted to base connections on the full range of PubMed publications of each authors, in order to identify collaborations outside of the meta-analyses under study. For this purpose, we used a code that, after downloading the full article list for each author name with initials as retrieved from a DOI or PMID search from PubMed itself, crosschecked each pair of authors within this article list, creating new connections or adding weight to existing ones according to the matched names if collaborations were found within the PubMed database (**Supp. Fig. 3**). However, after manually revising the retrieved articles for establishing author identity, we found that this method created a prohibitive number of spurious associations between researchers due to articles from homonyms (**Supp. Table 2**). Using author’s full names as retrieved from articles instead of initials as search seeds did not fully solve this problem. Thus, we chose to maintain the approach of using connections within the meta-analysis for the subsequent steps in order to prevent spurious clustering of unrelated authors. We note that, albeit infrequently, homonyms can also be an issue within a meta-analysis. Thus, we recommend to manually check for them among included studies before constructing co-authorship networks. Automated methods for author disambiguation have also been described in the literature^25^, but for individual meta-analyses manual screening is likely to be sufficient.

### Modularity analysis

To define author clusters, we used Gephi 0.9.2 to perform modularity analysis of author networks. We used the software’s default settings (i.e.: random decomposition; using weights from edges; resolution = 1), which uses the Louvain method for community detection^26^. After separation of authors into clusters, we manually assigned results from studies to their respective clusters. If a study had authors from different clusters, its results were assigned to the cluster with the most authors in the study. In the case of a tie (something that did not happen in our examples), effect sizes can be attributed to both groups, halving the sample size in each of them so as not to distort the meta-analytic effect estimate; alternatively, they can also be attributed to a separate cluster. As described previously, if a DOI did not retrieve any authors from PubMed, the results from this study became a cluster by itself. In our sample of meta-analyses, it only occurred in Mathie et al., where six studies included in the meta-analysis did not have any DOIs or PMIDs. The obtained clusters were used to build the collaboration networks in **Fig. 2** and the histograms showing the distribution of results among articles and clusters in **Fig. 3**.

**Figure 3.**
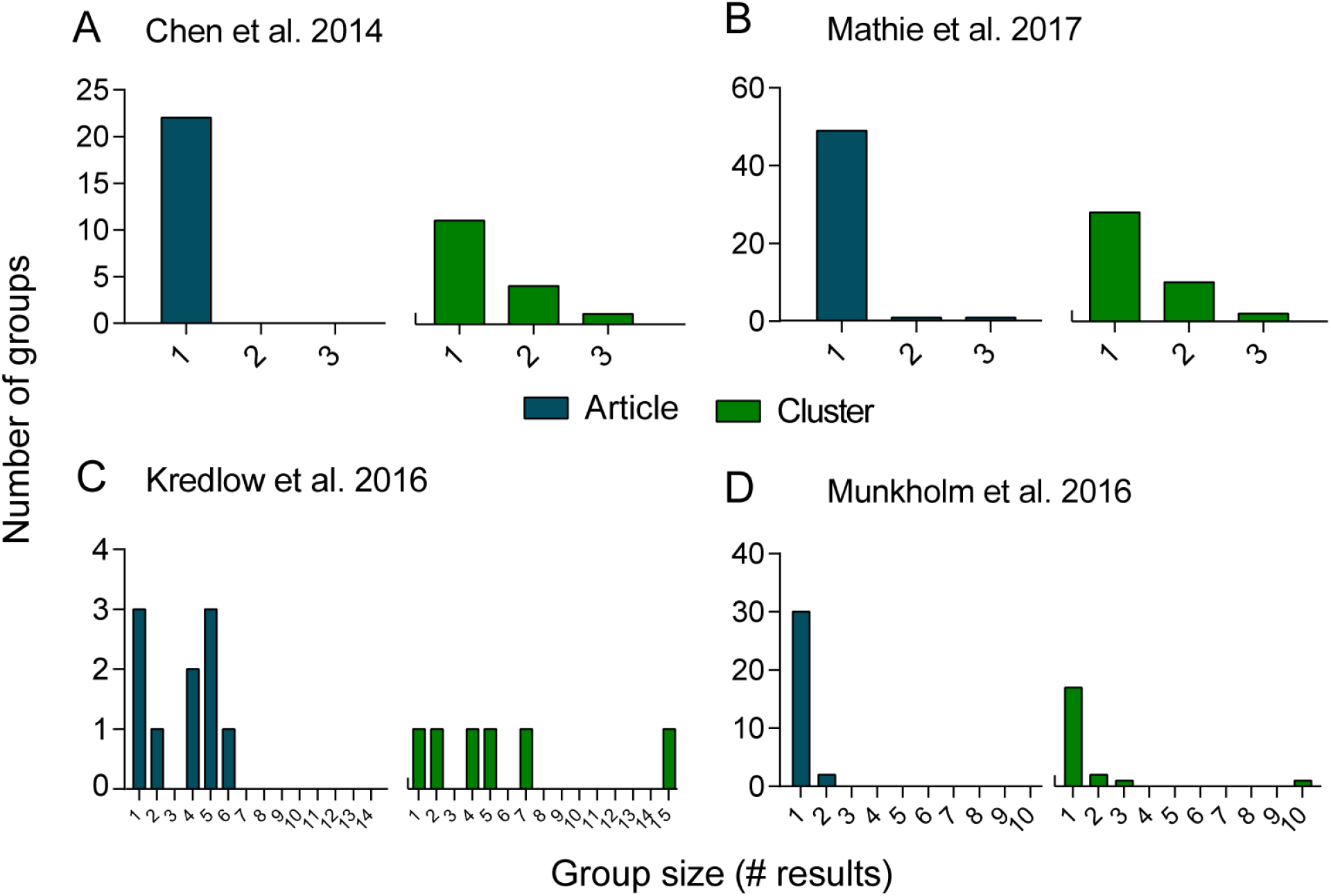
Distribution of results within meta-analyses. Histograms show how the results in each meta-analysis are aggregated when articles or author clusters are used as subgroups. Each bar represents a particular subgroup size (X axis), while the Y axis represents the number of subgroups of that size within the meta-analysis when results are grouped by article (left side, blue bars) or cluster (right side, green bars **(A)** Chen et al., 2014. Article: n=22, size=1; Cluster: n=16, size=1 to 3. **(B)** Mathie et al., 2017. Article: n=51, size=1 to 3]; Cluster: n=40, size=1 to 3. **(C)** Kredlow et al, 2016. n=10; size=1 to 6, Cluster: n=6, size= 1 to 15. **(D)** Munkholm et al., 2016. Article: n=32, size=1 to 2; Cluster: n=21, size=1 to 10.

### R^2^ estimation for articles and clusters

Data extracted from meta-analyses (effect size, sample size and standard error) was fed into Comprehensive Meta-Analysis version 3.3 (CMA, Biostat Inc.), which computed point estimates and variances for the studies. We then calculated the proportion of the variance explained either by (a) articles or (b) author clusters using two separate mixed-effects meta-regression models, using restricted-maximum likelihood (REML) estimator, with either the article or cluster as a categorical moderator (**Table 2**).

**Table 2.** R^2^ calculations for article and author cluster groups. After grouping study results according to either cluster or article membership, we computed the amount of the total between-results variance explained by subgroup membership using a mixed-effects meta-regression model using either cluster or article as a moderator. The fraction of the variance accounted for by the subgroup is expressed as R^2^. For each R^2^, a p-value was calculated by a bootstrap method based on 1,000 reshufflings of the results within each meta-analysis structure, maintaining the same number of articles or clusters (see Supp. Fig. 4).

| Reference | Grouping (n) | R<sup>2</sup> | p-value |
| --- | --- | --- | --- |
| Chen et al. 2014 <sup>[18]</sup> | Article (22) | - | - |
|  | Cluster (16) | 0 | 0.136 |
| Mathie et al. 2017 <sup>[19]</sup> | Article (51) | 0 | 0.609 |
|  | Cluster (40) | 0 | 0.146 |
| Kredlow et al. 2016 <sup>[20]</sup> | Article (10) | 0.828 | <0.001 |
|  | Cluster (6) | 0.691 | <0.001 |
| Munkholm et al. 2016 <sup>[21]</sup> | Article (32) | 0 | 0.622 |
|  | Cluster (21) | 0.311 | 0.017 |

The computation of R^2^ follows the equation 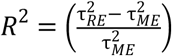, where 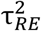 is the total amount of heterogeneity, as estimated based on a standard random-effects model, and 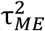 is the amount of residual heterogeneity, as estimated based on the mixed-effects meta-regression model. Using this calculation, the sampling distribution of R^2^ values is dependent on the number and size of subgroups (i.e. the distribution of individual results among articles or clusters): the fewer and smaller the subgroups, the greater the chance of finding spurious high values of R^2^ by chance alone, due to inaccurate estimation of variance within them. As the grouping structure varied widely across meta-analyses, we used a bootstrapping method for inferring the p-value. We constructed an R code to perform a Monte Carlo permutation test, randomly reshuffling study results within each meta-analysis 1000 times, while maintaining its structure in terms of number and size of subgroups. We then estimated p-values for the R^2^ values found in each meta-analysis by calculating their correspondent percentile in the generated R^2^ probability density distributions (**Supp. Fig. 4**). The files for all meta-analyses in CMA and the R codes for p-value calculations are available as supplementary material.

### Detecting deviant author clusters

To detect research groups with results differing from the rest of the literature (**Figs. 4-7**), we used the R package metafor^27^ to compare the estimates of each author cluster with the meta-analytical estimate of the remaining studies. For each comparison, we assumed that the cluster and the remaining studies each represented an independent random-effects model and calculated the estimate and standard error for both, using the REML estimator for τ^2^. We then combined these two estimates in a fixed-effects model, using these two estimates as a moderator and testing for its significance using a Wald-type test of the difference between the two estimates. This approach is similar to a multiple random-effects meta-regression, as both have similar performance in terms of Type I error rates and statistical power, and it is preferable when residual between-studies variances are clearly different^28,29^. We adjusted all p-values for the number of tests conducted within each meta-analysis using a Bonferroni-equivalent p-value correction. The R codes for these comparisons are also available as supplementary material. Finally, it is important to notice a distinction from this method to the traditional sensitivity analysis, in which a meta-analysis is compared with and without a given result. Instead, our method compares the cluster itself with the meta-analysis without it.

**Figure 4.**
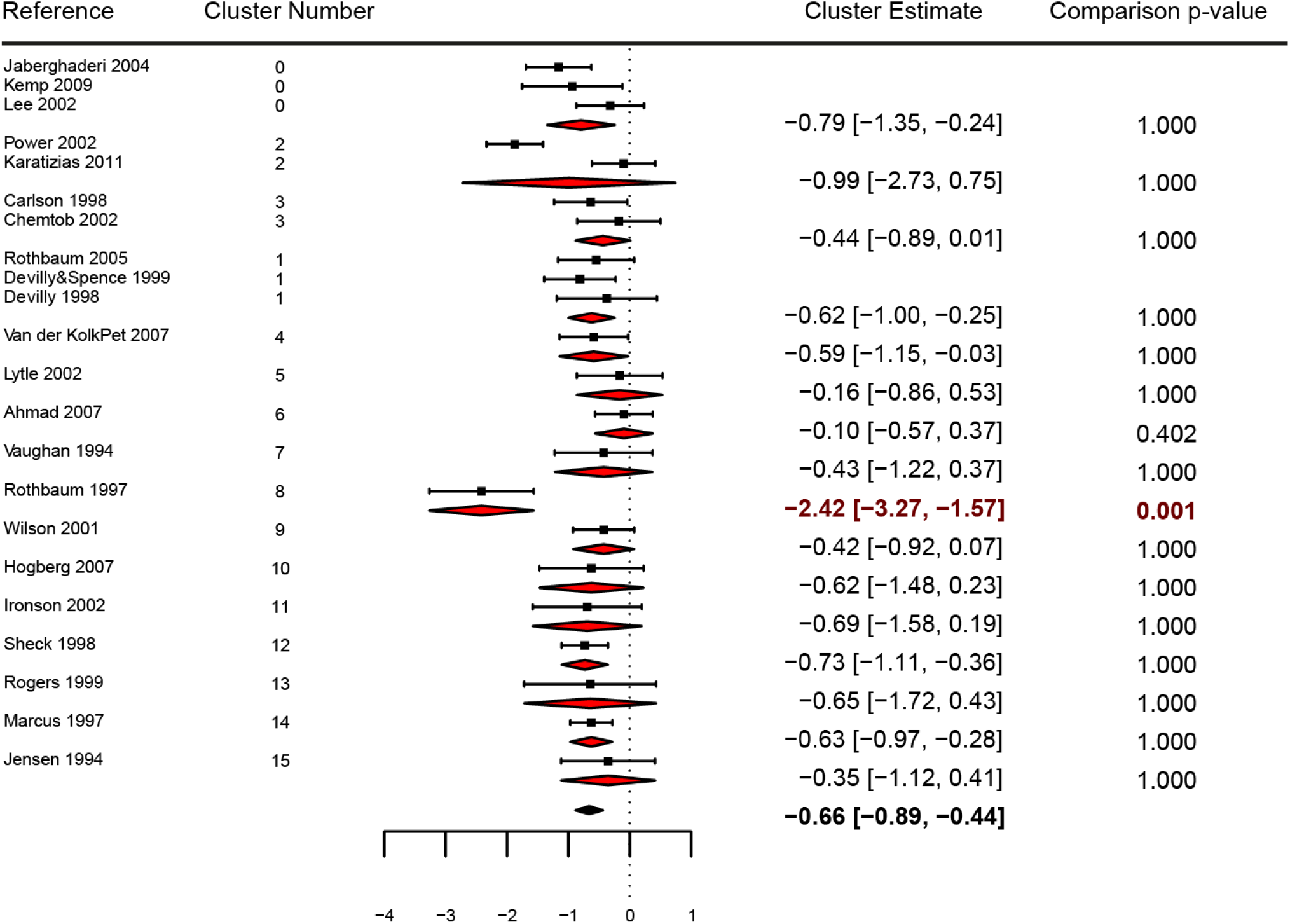
Forest plot of Chen et al., 2014. The plot shows the effect size (in Hedges’ g) and error (95% CI) of individual studies (squares), the estimates of meta-analyses for each author cluster (red diamonds) and the overall estimate (black diamond) using standard random-effects models. Each subgroup was compared against the remaining studies within the meta-analysis by a Wald-type test, yielding Bonferroni-corrected p-values shown on the right column. Estimates and corrected p-values of clusters significantly differing from the rest of the meta-analysis at an α of 0.05 are shown in red.

**Figure 5.**
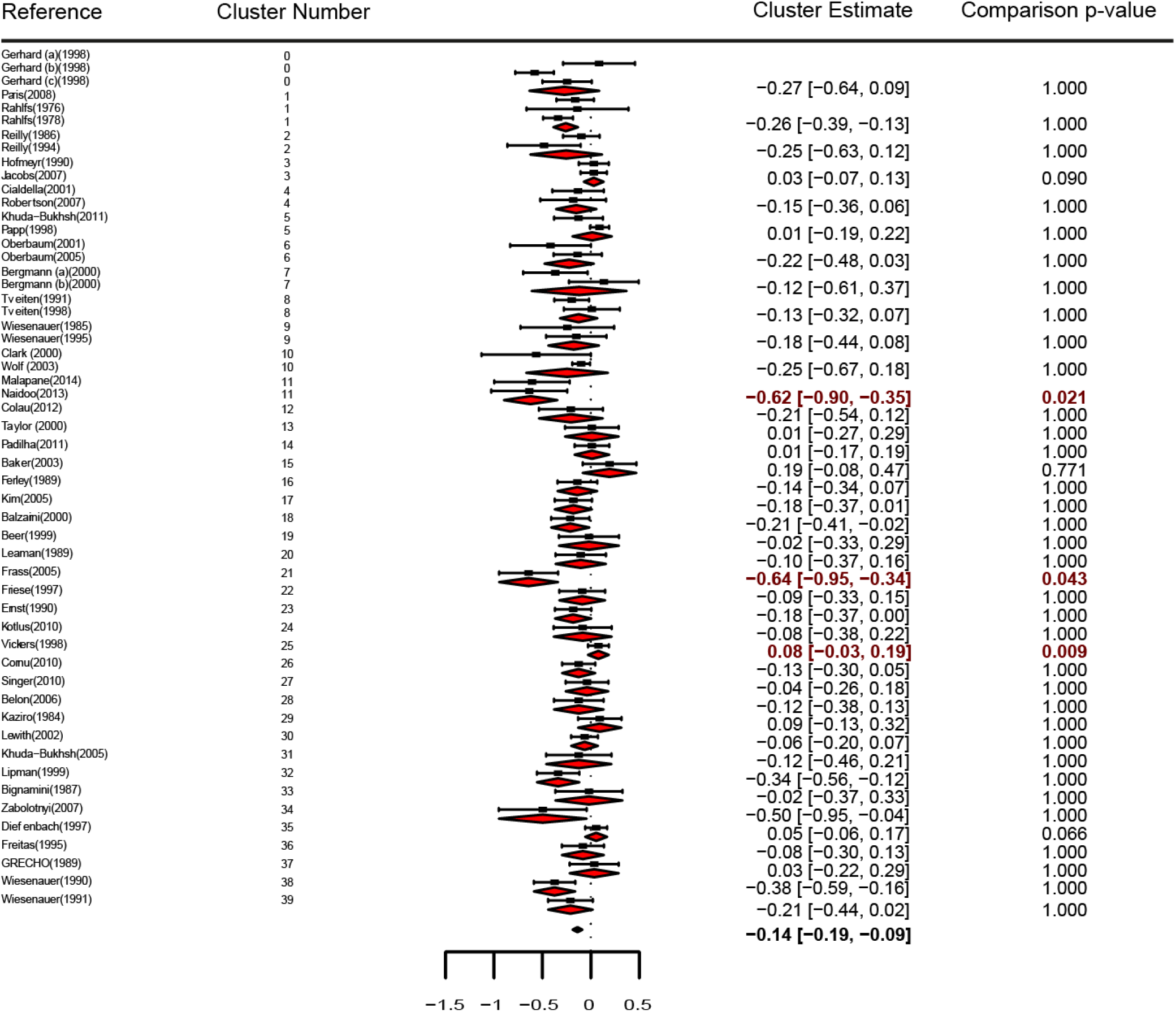
Forest plot of Mathie et al., 2017. Effect size, error and estimates are represented as in Figure 4.

**Figure 6.**
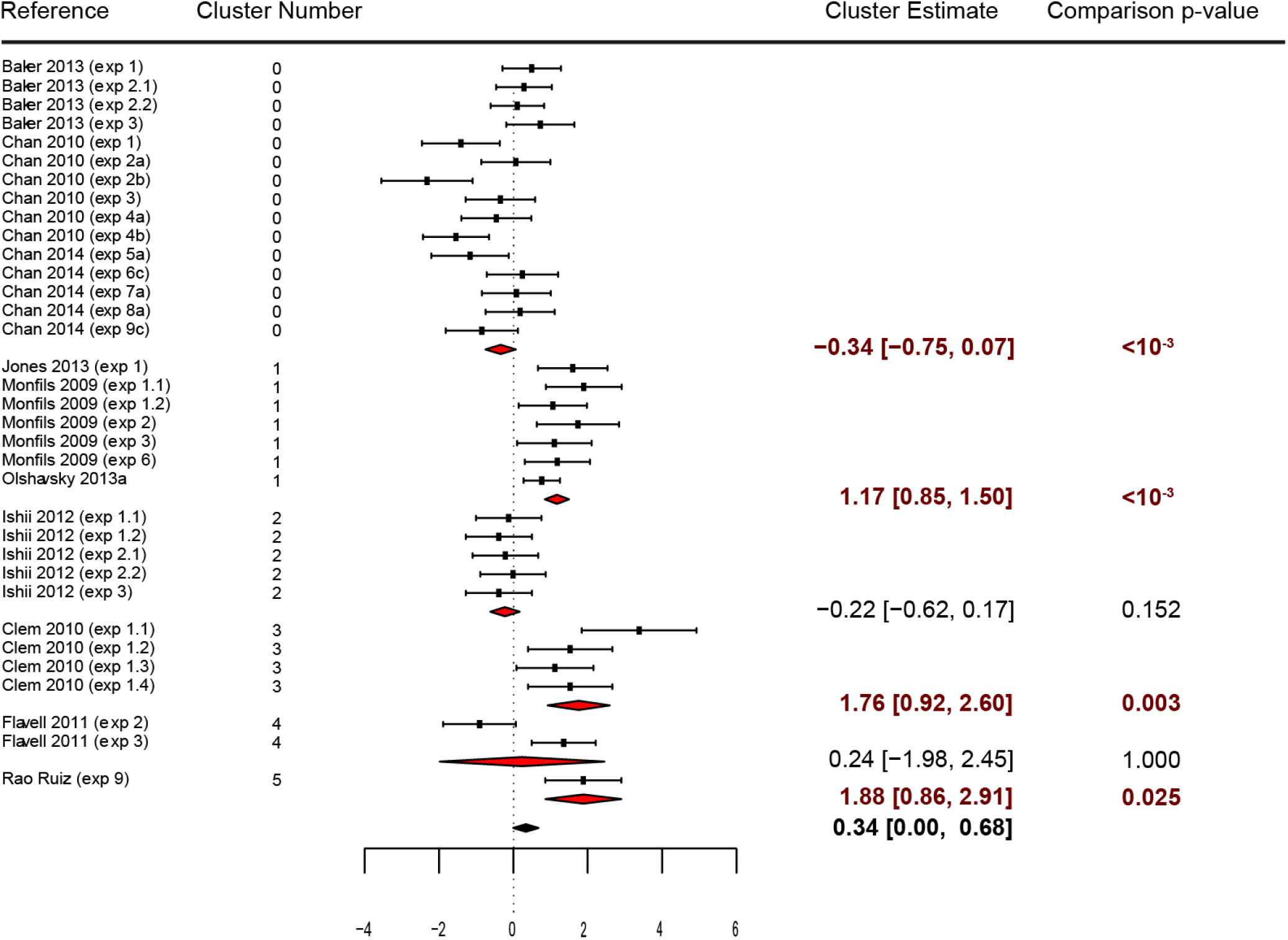
Forest plot of Kredlow et al., 2017. Effect size, error and estimates are represented as in Figure 4.

**Figure 7.**
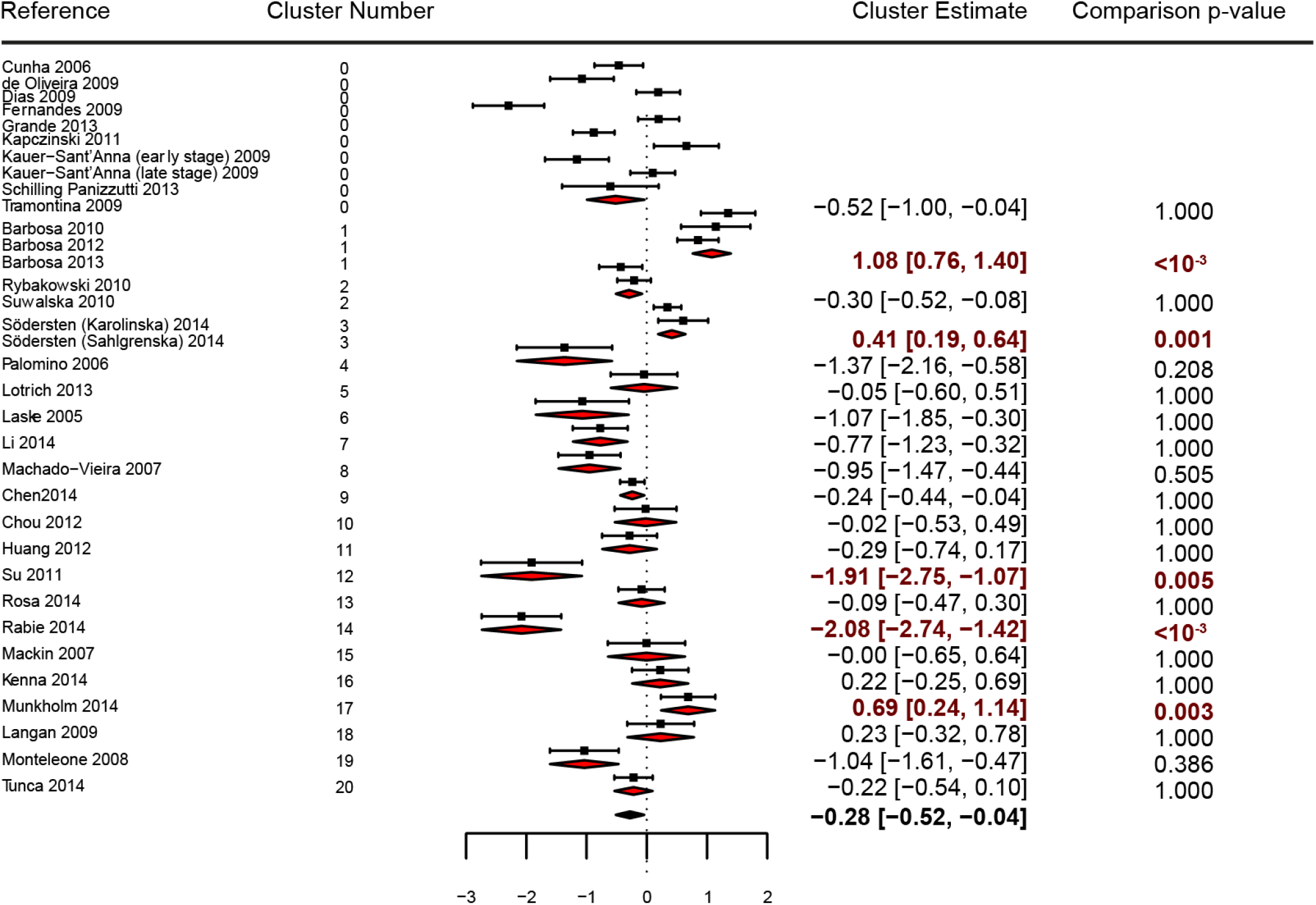
Forest plot of Munkholm et al., 2017. Effect size, error and estimates are represented as in Figure 4.

### Correction of estimates by multilevel analysis

After clustering results from the meta-analyses, the effect size estimates obtained across studies are nested within two higher-level grouping variables (i.e. article and research group), whose impact on heterogeneity can get tangled up when they are analyzed separately. Moreover, unbalanced representations between different articles or research groups can bias meta-analytic estimates towards the effects found by a highly-represented research group, making them less representative of the literature as a whole. To control for this, we used the metafor R package to employ the multilevel meta-analytic model described by Konstantopoulos^15^. We calculated the overall estimate and variance components for this multilevel model, adding random effects both at the level of articles and author clusters (**Supp. Table. 3**). This analysis, as well as our previous R^2^ calculation (**Table 2**), demonstrated that the article level had negligible influence on three of the four meta-analyses; moreover, since these had few articles with more than one results, the multi-level model runs the risk of being distorted by inaccurate estimation of within-article variances. Thus, for our main multilevel analysis, we considered only the author cluster level. We then compared these results with those obtained with a standard random-effects model that did not consider the group of origin. (**Table 3**). A forest plot showing article-level estimates for Kredlow et al. 2016 is shown in **Supp. Fig. 5**. The codes for these analyses are provided as supplementary material.

**Table 3.** Standard random-effects model and multilevel random-effects model analyses. The standard two-level model does not take author cluster into consideration, while the multilevel model uses this a level (with results nested within author clusters). The table shows the effect estimate, 95% confidence interval and p-value for both models, the overall between-study variance component τ^2^ for the standard two-level model and the separate within-level variance components (σ^2^) for author cluster and individual results in the multilevel analysis.

|  | Reference | Chen et al.<sup>[18]</sup> | Mathie et al.<sup>[19]</sup> | Kredlow et al.<sup>[20]</sup> | Munkholm et al.<sup>[21]</sup> |
| --- | --- | --- | --- | --- | --- |
| Standard analysis | **Estimate** | -0.66 [-0.89,-0.44] | -0.14 [-0.19,-0.09] | 0.34 [-0.03, 0.71] | -0.29 [-0.58,-0.00] |
|  | **p-value** | <0.001 | <0.001 | 0.072 | 0.047 |
| | $\tau^2$ | 0.187 [0.07, 0.50] | 0.019 [0.01, 0.04] | 0.987 [0.58, 2.83] | 0.656 [0.41, 1.24] |
| Multilevel analysis | **Estimate** | -0.66 [-0.89,-0.44] | -0.14 [-0.19,-0.09] | 0.69 [-0.09, 1.48] | -0.31 [-0.66, 0.04] |
|  | **p-value** | <0.001 | <0.001 | 0.084 | 0.080 |
| | $\sigma^2_{\text{cluster}}$ | 0.000 [0.00, 0.18] | 0.004 [0.00, 0.03] | 0.797 [0.20, 4.43] | 0.278 [0.00, 0.92] |
| | $\sigma^2_{\text{result}}$ | 0.187 [0.07, 0.47] | 0.015 [0.00, 0.04] | 0.290 [0.09, 0.71] | 0.397 [0.18, 0.90] |

### Supplementary material

All codes and files necessary to the analyses mentioned in this work are available at the Synapse Repository at https://doi.org/10.7303/syn21438273

## Results

### Meta-analysis features

We initially extracted data from the four meta-analyses to use as case studies. Two of them^18,19^ were of clinical intervention studies, one concerned behavioral studies in rodents^20^, and the other comprised biomarker studies in patients^21^. There was significant heterogeneity in all four, as reflected by Q-tests and I^2^ values (**Table 1**). Egger’s regression indicated small-study effects suggestive of publication bias for two of the meta-analyses, but only one had a high number of missing studies according to trim-and-fill analysis, as shown by the funnel plots in **Supp. Fig. 1**.

### Defining research groups by collaboration networks

To define research groups, we constructed graph networks using individual study authors in each meta-analysis as nodes, with the weights of edges defined by the number of studies coauthored within the meta-analysis. Modularity analysis separated these authors into clusters representing research groups, represented in different colors in **Fig. 2**.

Histogram distributions for the number of results per article and research group (**Fig. 3**) show that the majority of clinical studies had a single result per article. On the other hand, in the meta-analysis of rodent studies by Kredlow et al., a much higher number of results per article is found. After aggregating results by author cluster in Chen et al. and Mathie et al., we could identify only a few groups with more than one study, and none with more than three. On the other hand, after applying the same procedure in Kredlow et al. and Munkholm et al, we observed the appearance of author clusters contributing up to 15 results.

### Influence of article and author cluster of origin on effect sizes

To measure how much of the heterogeneity in each meta-analysis could be attributed to the author cluster and/or to the article of origin, we calculated the amount of the total between-results variance that could be explained by grouping the results according to either cluster or article membership (**Table 2**). In meta-analyses where most articles contributed a single result, the article of origin explained none of the overall heterogeneity. However, in Kredlow et al., the article of origin explained most of the observed heterogeneity across experiments, an influence that was also captured at the author cluster level. For Chen et al. and Mathie et al., there was no statistically significant influence of authorship on the variance. In Munkholm et al., on the other hand, the amount of heterogeneity explained by authorship was smaller, but statistically more significant.

### Detecting deviant author clusters

The approach described above allowed us to quantify the influence of authorship on heterogeneity, but not to attribute this effect to specific author clusters. In order to do that, we compared the effect estimates of each author cluster with that of the remaining studies within the meta-analysis. When applying this method to Chen et al. (**Fig. 4**) and Mathie et al. (**Fig. 5**), in which the authorship effect is small, just one cluster out of 16 (6.2%) in the former and 3 out of 40 (7.5%) in the latter were significantly different from the rest of the results after controlling for multiple comparisons. Conversely, in Kredlow et al. (**Fig. 6**), where the number of clusters was smaller and there was a high impact of authorship on heterogeneity, 4 out of 6 clusters (66.7%) were significantly different from a meta-analysis excluding their own results. In Munkholm et al. (**Fig. 7**), there was also evidence of authorship bias, with 5 of 21 clusters (23.8%) differing significantly from the rest of the meta-analysis.

### Correcting effect estimates by multilevel analysis

After quantifying the amount of variance attributable to articles and author clusters, we used random-effects multilevel models using either cluster (Chen et al., Munkholm et al., Mathie et al.) or both cluster and article (Kredlow et al.) as nested levels to summarize the results. This allowed us to correct for the effects of non-independence on effect size estimates, as well as to differentiate the effects of article and author cluster membership when both were present. **Table 3** shows the estimates obtained with these models, comparing them to a standard random-effects model that does not take authorship into account.

For meta-analyses with no significant authorship effect on heterogeneity (Chen et al. and Mathie et al.), the multilevel model showed negligible influences of the cluster-level component of heterogeneity, leading to effect estimates that were almost identical to those of the standard two-level model. On the other hand, for Kredlow et al., in which strong evidence of authorship bias was found, we observed a twofold change in the estimate of the multilevel model when compared to the standard one. For this meta-analysis, multilevel modelling showed that variance was explained both by the cluster and article levels, with a higher value for the cluster component (**Supp. Table 3**), as can be observed in a forest plot with estimates grouped by article (**Supp. Fig. 5**). Lastly, for Munkholm et al., the cluster component maintained its effect on heterogeneity, slightly changing the multilevel model effect estimate and leading to a wider confidence interval and a higher p-value than the standard model.

## Discussion

Meta-analyses and systematic reviews have been used for decades to synthesize scientific data, shaping evidence-based policies, and guiding medical decisions^31^. For these summaries to be reliable, however, meta-analyses should not simply summarize the literature, but also help to identify biases and other pitfalls in order to correct for them^32^. Many of these methods are used routinely nowadays, such as Egger’s regression, funnel plots^33^, and trim-and-fill analysis^34^ to detect small study effects suggesting publication bias, I^2^ calculations to evaluate heterogeneity^35^, and excess significance tests^36^ to detect preferential reporting of significant findings and/or p-hacking.

In this work, we describe a simple method to detect and correct for authorship bias in meta-analyses. This phenomenon happens when results from the same laboratory or research group are summarized without proper correction for non-independence, potentially giving excessive weight to results from a single group in estimate calculations. This type of bias has mostly gone unattended in the available literature, perhaps because most clinical meta-analyses are performed based on a small number of studies, usually containing a single result each^37^.

Isolated evidence has suggested the presence of authorship bias in specific fields of research. For instance, in a meta-analysis of violence risk assessment tools, it was shown that tool designers found more positive results than independent investigators evaluating other researchers’ tools^38^. A recent meta-regression study on randomized trials on the safety of hydroxyethyl starch also identified that a specific research group, with a history of retractions due to data manipulation, had significantly different effect sizes when compared to other groups^39^. Nevertheless, these investigations have been carried out on an individual basis, using different methodologies in each case. We believe that having a standard method for automatically attributing authorship to different groups can allow this kind of analysis to be performed more systematically in meta-analyses.

The problem of non-independence among results is much more marked in meta-analyses from preclinical studies, which have been on the rise in recent years^40^. These types of studies often have smaller sample sizes and greater heterogeneity among them than clinical studies; moreover, each article frequently contributes with several different experiments to the same meta-analysis^40,41^. Thus, it is not uncommon for a single lab to account for a large fraction of the research in a given area. Accordingly, in our example of a preclinical meta-analysis^20^, we identified a strong influence of both the article and research group of origin on effect sizes. We believe that this kind of non-independence may be the rule for meta-analyses of non-human biomedical research; thus, tools that can detect and account for this phenomenon can be especially useful in this field.

The main contribution of our method is to provide an objective, unbiased definition of a research group. This definition is usually highly subjective, as group affiliation and collaboration patterns are variable and dynamic. We have circumvented this issue by creating a collaboration network graph based on the meta-analysis itself and using modularity algorithms to detect author communities within it. This method is based on collaboration between researchers – thus, even scientists who are not currently in the same research group or laboratory can be aggregated if they are highly collaborative. We believe that this method can capture groups of researchers with similar views, methodological preferences and interpretations, and thus provide an objective, data-driven approach to detect authorship bias. The fact that authorship influence was detected in 2 out of 4 meta-analyses evaluated in our study shows that this form of clustering captures real sources of heterogeneity, and provides initial validation of our method as a useful tool for further analyses of the literature.

An arguably intuitive option for our method would be adding a moderator variable that reflects the place where the study was conducted; however, we believe that there are several limitations in this approach. First, an article with authors from different institutions likely means that experiments took place in different locations. In this case, one should make a choice on which is the main place for that investigation (e.g.: by the affiliation of the corresponding author, or by the number of authors related to one location), adding subjectivity to the analysis. Second, it is somewhat common that a researcher develops a body of literature in one institution but later move places, maintaining the same methods and analyses from the former laboratory. Third, there is the case of multicenter clinical studies, where patients of diverse locations across the globe provide samples for analysis in a single laboratory, which could even be different from the research group that designed the study. Thus, the idea behind the cluster analysis is precisely to provide an objective, data-driven approach to define authorship communities. Moreover, although it is probable that the authorship cluster will occasionally reflect the laboratory location, we believe that in most settings it would be more useful than adding a location-based moderator, as the latter ignores the influence of authors’ experimental and analytical handling.

Nevertheless, our method of creating graphs has limitations. As it was fully based on co-authorship within the studies included in the meta-analysis, it is likely that many collaborations will go undetected, as authors can work together in articles outside of this sample. We attempted to avoid this issue and improve our detection of collaborations by using PubMed searches of single authors in order to construct lifetime collaboration graphs (**Supp. Fig. 3**). However, the sheer lack of specificity of names and initials – which are still the seeds for most database searches in science – generated a prohibitive amount of false-positive collaborations that distorted the resulting graphs (**Supp. Table 2**). As unique author identifiers such as ORCID^42^ become more popular, however, it is likely that such approaches will be more feasible in the near future – and in that case, lifetime collaborations might ultimately yield better authorship maps than individual meta-analyses.

A simple tool such as ours might plausibly be incorporated in meta-analysis packages to provide a simple assessment of authorship bias. Although it currently runs on partly on proprietary software (i.e. MATLAB), similar implementations can be obtained using other platforms – a preliminary analysis shows that using VOSViewer, a tool for constructing bibliometric networks^24^, leads to very similar results (**Supp. Fig. 2**). The clustering algorithm itself is built with open-source software (Gephi) and based on well-known mathematical algorithms for dealing with graph clustering^26^. Thus, although our initial implementation and validation of the tool has been performed on different software platforms, a plausible short-term development is to incorporate these different functions within a unified package.

In this work, we have focused on the immediate advantages of detecting authorship bias within an individual meta-analysis. After detecting and quantifying the percentage of heterogeneity due to authorship, we showed that this effect could be attributed to individual clusters in some meta-analyses. This resembles sensitivity analysis, a procedure that is routinely performed in meta-analyses^43^, but is based on groups rather than individual results, thus providing a way to detect research groups yielding results that deviate from the remaining ones. The interpretation of these discrepant results can vary, but an objective way to prevent the output of a single research group from inappropriately distorting meta-analytical estimates is to perform multilevel modeling based on author clusters. In our work, we show that this approach can have a large effect on individual estimates, especially in situations with high clustering of results, as in the case of preclinical research.

Although we have referred to the effect of authorship on effect sizes as ‘authorship bias’, it should be clear that such bias is not necessarily due to authors’ perceptions and beliefs. There are myriad sources of variability that can occur due to methodological choices that, if consistent within a research group, can lead to bias towards smaller or larger effects. Studies of inter-laboratory variability in basic science have shown that, even when careful measures are taken to ensure methodological homogeneity, a large amount of the variance among experiments is attributable to the laboratory where they are performed^44,45^. The same is true for clinical populations, which are likely to be more similar within the work of a single research group than across groups. Meta-regression of specific methodological variables within studies can help to assess whether these variables can account for the effect of authorship; nevertheless, even if no such moderators are found, one cannot rule out the possibility that unassessed methodological factors can be responsible for variability in results among research groups.

Finally, although our work was focused on the application of authorship clusters to provide insights on the meta-analyses themselves (e.g. effect estimate correction and detection of deviant groups), a tool for evaluating authorship bias can also have more widespread applications in understanding how authorship influences results in different fields of science. Although our limited sample does not allow us to generalize our conclusions, it is interesting to note that the impact of authorship on effect sizes was very different between meta-analyses of clinical and preclinical data. Whether these and other patterns of authorship bias hold true in larger, representative samples of meta-analyses from different fields of research is an open question that tools such as ours can help to tackle, providing wider insights on the interactions between authorship and study results.

## Data Availability

Both our data and code are available as supplementary material, along with instructions for using the code to construct the figures of our article.

## Acknowledgements

The authors are indebted to Roberto Maia for participating in data extraction, to Giovanni Salum for help with coding in R, and to Maria Alexandra Kredlow for sharing meta-analysis data.

## Highlights

What is already known:

- Meta-analyses are prone to various types of bias that can influence effect size estimations;
- One of the source of bias in meta-analyses is non-independence between study results from the same research group;

What is new:

- We develop a method to automatically define research groups, by creating a collaboration network graph based on studies in a meta-analysis and using modularity algorithms to detect author communities;
- We demonstrate that the research group of origin can impact effect size in distinct types of meta-analyses;
- We show that multilevel random-effects meta-analytic models can be used to correct this type of bias.

Potential impact for RSM readers outside the authors’ field

- The described method can be applied to any kind of meta-analysis to estimate authorship bias, regardless of research field;

---

**Supplementary Table 1.**
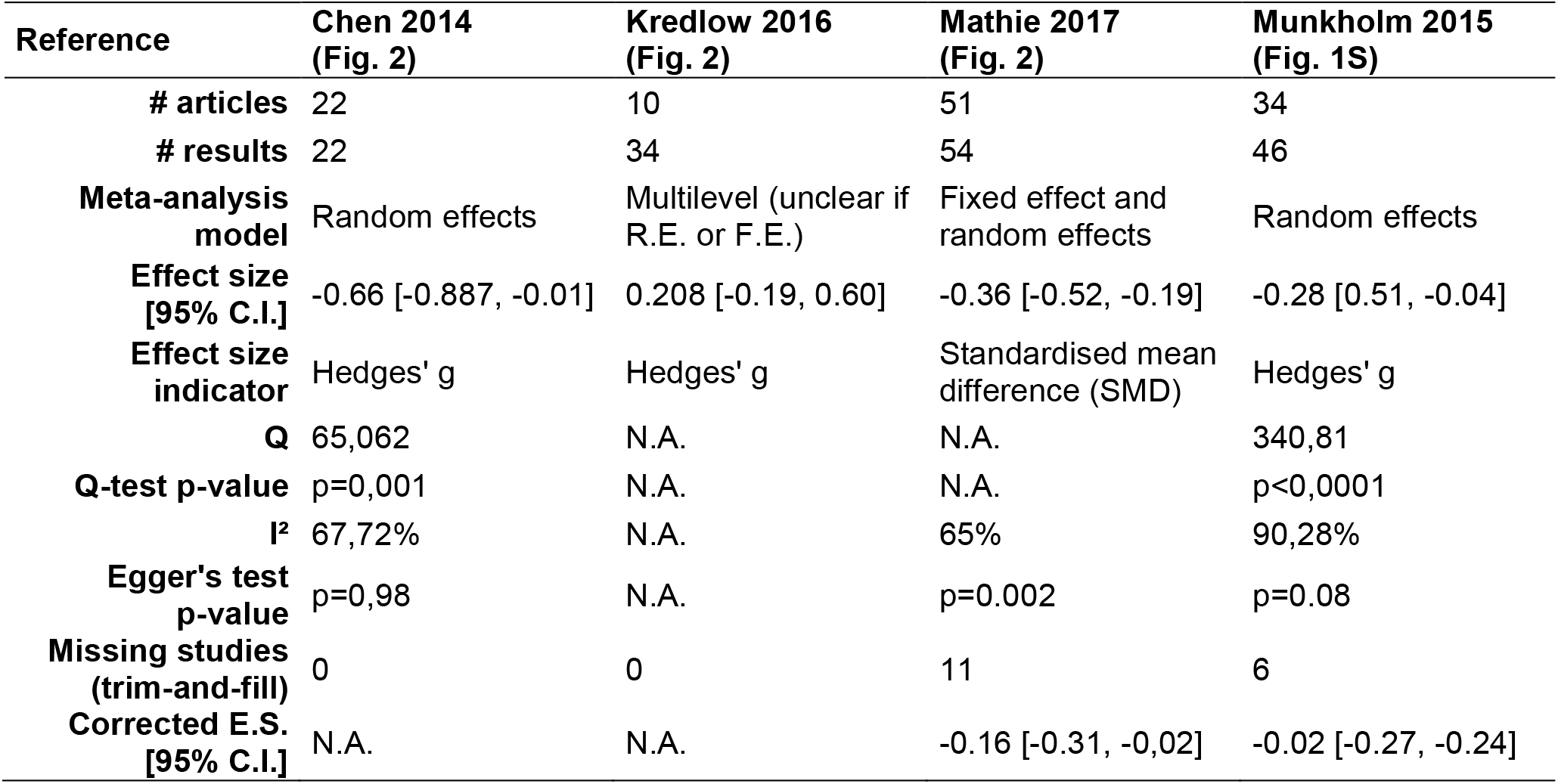
Meta-analysis features from original articles. The analytic models, computed effect sizes, indicators of heterogeneity (Q-test p values and I^2^ values), and small-study effects (Egger’s regression p-values, as the number of missing studies in trim-and-fill analysis) were retrieved from the descriptions in the original articles.

**Supplementary Table 2.** Analysis of differences between networks built using different methods. For each method, we plot the number of (a) lost edges, (b) spurious edges, (c) edges with reduced weight due to lost papers, and (d) edges with increased weights due to spurious collaborations when each of the 3 automated methods shown on Supp. Fig. 3 is compared to a manually verified ‘gold standard’ network of lifetime collaborations. Lifetime DOI-based search for articles in PubMed using last names and initials added a significant number of new connections, leading to the spurious aggregation of clusters that were separated in the verified network. Full-name search on PubMed was able to reduce this effect, but still led to the formation of many spurious edges. As expected, within-meta-analysis searches did not detect many of the edges formed by lifetime collaboration graphs; on the other hand, it did not lead to the formation of any spurious connections.

|  | PubMed Initials | PubMed Full Name | Within Meta-Analysis |
| --- | --- | --- | --- |
| **Lost edges** | 4 | 3 | 24 |
| **Spurious edges** | 25 | 15 | 0 |
| **Decreased-weight edges** | 13 | 42 | 194 |
| **Increased-weight edges** | 35 | 32 | 0 |

**Supplementary Table 3.** Multilevel random-effects model analyses including article as a level. We applied a multilevel model considering both the author cluster and article as levels (with article nested within author cluster) for all meta-analyses. The table shows the effect estimate, 95% confidence interval and p-value for both models, the separate within-level variance components (σ^2^) for author cluster, article and individual results in the multilevel analysis.

| Reference | Chen et al. <sup>[18]</sup> | Mathie et al. <sup>[19]</sup> | Kredlow et al. <sup>[20]</sup> | Munkholm et al. <sup>[21]</sup> |
| --- | --- | --- | --- | --- |
| **Estimate** | -0.66 [-0.89,-0.44] | -0.14 [-0.19,-0.09] | 0.69 [-0.07, 1.45] | -0.31 [-0.66, 0.04] |
| **p-value** | <0.001 | <0.001 | 0.077 | 0.080 |
| $\sigma^2_{\text{cluster}}$ | 0.000 [0.00, 0.18] | 0.004 [0.00, 0.03] | 0.620 [0.00, 4.05] | 0.278 [0.00, 0.92] |
| $\sigma^2_{\text{article}}$ | 0.094 [0.00, 0.47] | 0.000 [0.00, 0.02] | 0.227 [0.00, 1.55] | 0.000 [0.00, 0.49] |
| $\sigma^2_{\text{results}}$ | 0.094 [0.00, 0.47] | 0.015 [0.00, 0.04] | 0.143 [0.00, 0.69] | 0.397 [0.00, 0.90] |

**Supplementary Figure 1.**
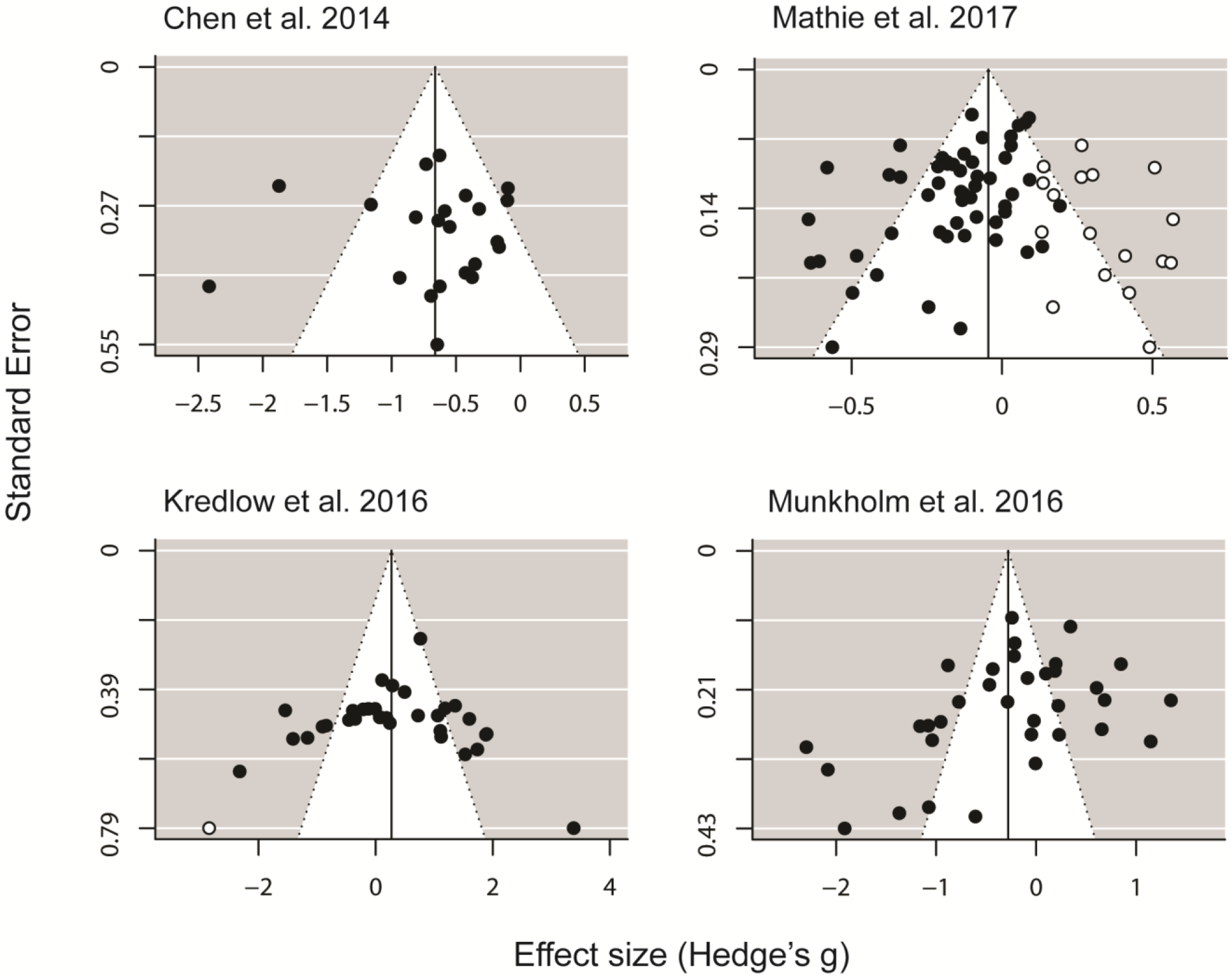
Funnel plots of included meta-analyses. After data extraction from the original meta-analyses, funnel plots were built using the metafor R package. Vertical lines represent the uncorrected meta-analytical estimates, black dots represent existing studies and white dots represent missing studies according to trim-and-fill assessment.

**Supplementary Figure 2.**
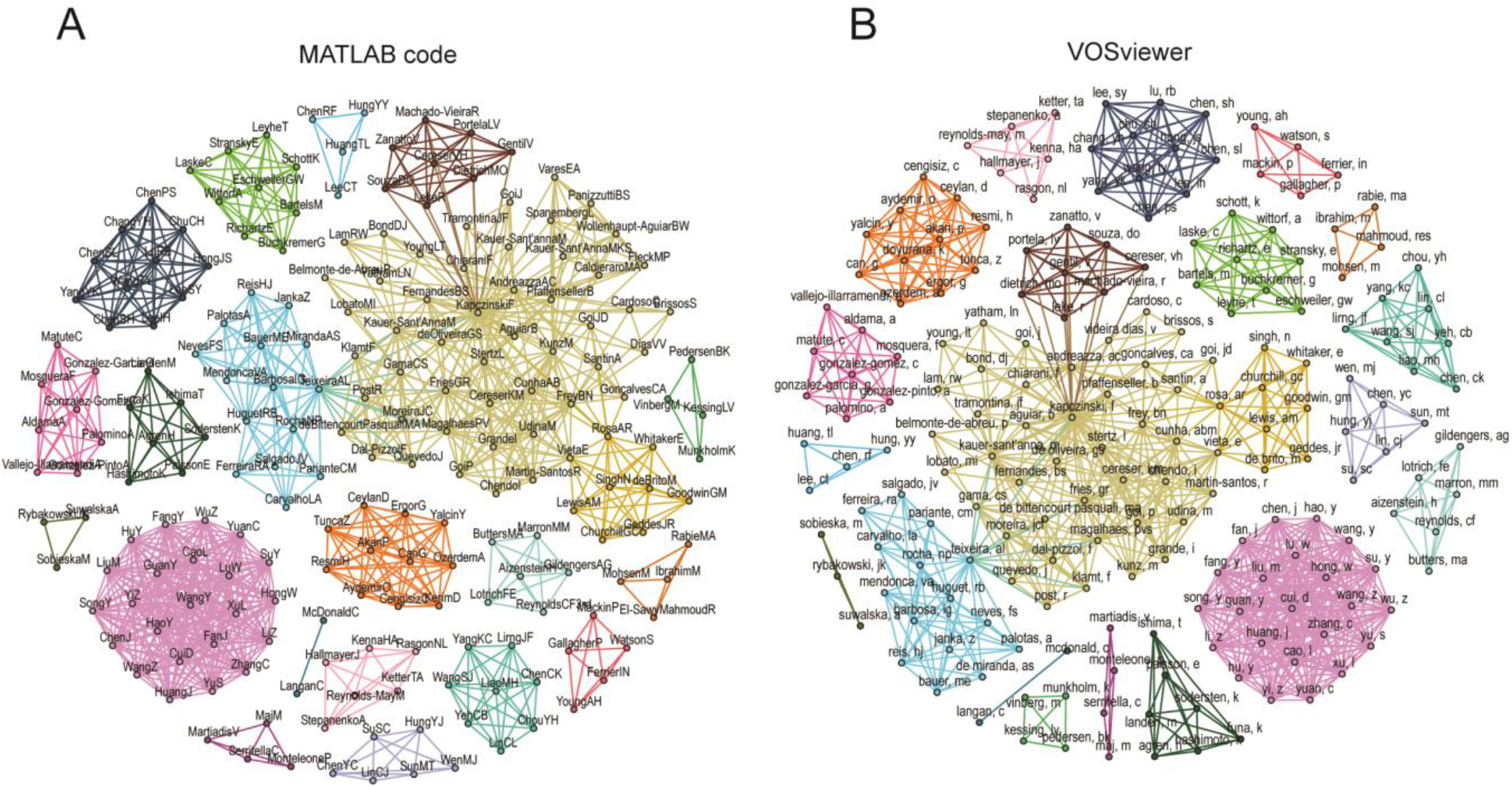
Author network generation using different software methods. **(A)** Author network in the Munkholm et al., 2016 meta-analysis (Fig. 1S in the original paper) built using MATLAB code: Network has 202 nodes, 1010 edges, 21 clusters; **(B)** Same network built using VOSviewer: Network has 194 nodes, 964 edges, 21 clusters. Although the first method identified more nodes (and consequently more edges), the overall network structure was similar, with the same number of author clusters. Equivalent clusters are similarly colored, although cluster positions are randomly assigned.

**Supplementary Figure 3.**
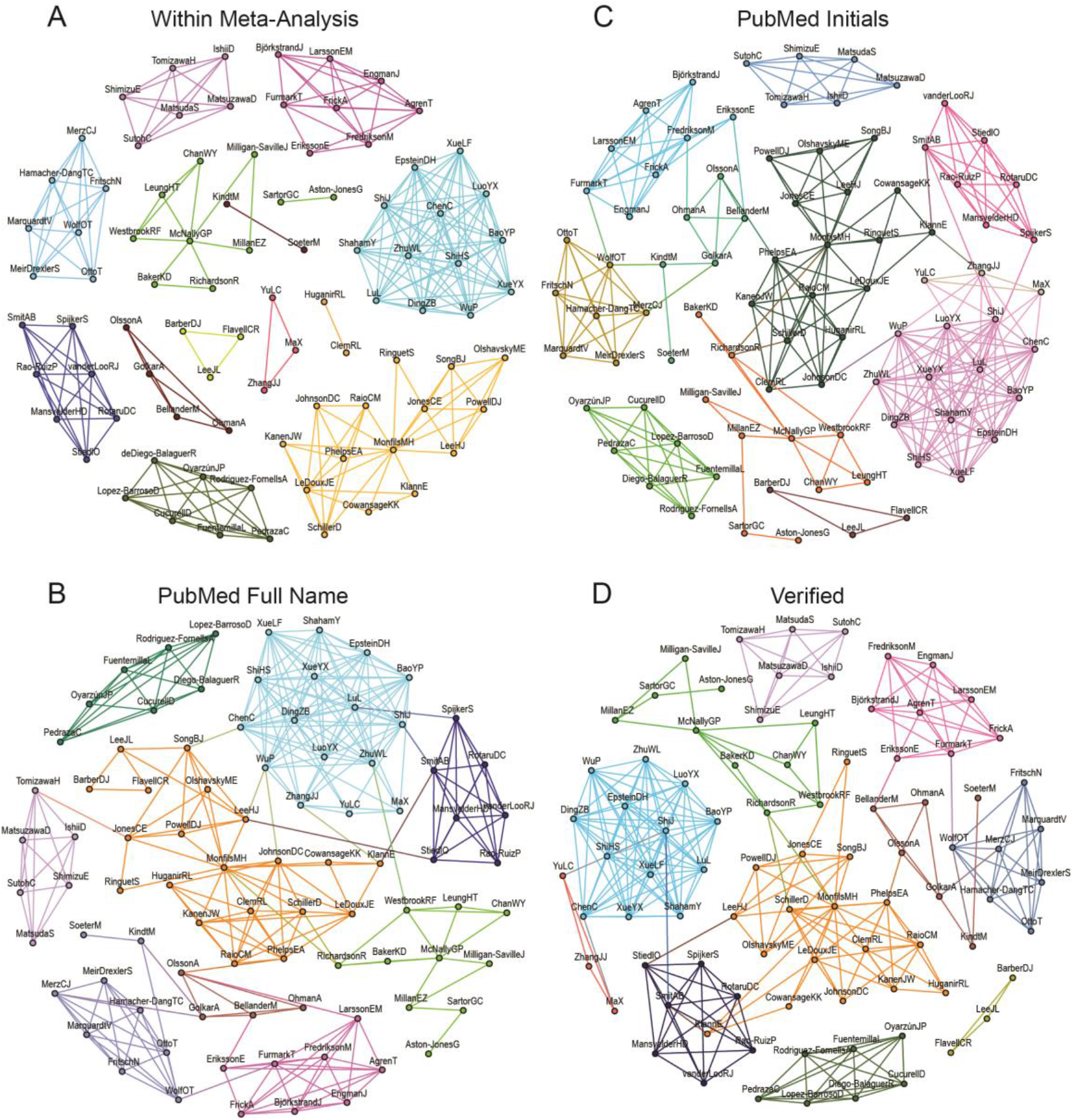
Comparison between distinct methods for identifying collaborations. We built networks using four different approaches to detect co-authorships between authors within all three meta-analyses included in Kredlow et al., 2016: **(A)** Within-meta-analysis connections, where authors are connected only by articles included in the meta-analysis itself. Notice that this network is different from that shown in Figure 2, which refers to a specific meta-analysis within the study. Network contains 87 nodes, 248 edges and 14 clusters; **(B)** Lifetime DOI search, where we used DOI numbers to retrieve researchers’ abbreviated names from PubMed, which were then used to search lifetime publications for each author. Based on these lists, we identified connections between authors by automated name crosschecking. Network contains 87 nodes, 293 edges and 9 clusters. **(C)** Lifetime full name search, where we manually retrieved authors’ full names (i.e.: non-abbreviated first names) from studies included in the meta-analyses and used these as search seeds to retrieve lifetime publications in PubMed and identify collaborations, in an attempt to minimize spurious hits caused by homonyms. Network contains 87 nodes, 284 edges, 11 clusters. **(D)** Lifetime verified connections, where we searched each pair of researchers using abbreviated names in PubMed, and then manually verified each article in the output to exclude spurious connections. Network contains 87 nodes, 272 edges, 11 clusters. For all graphs, cluster colors and positions are randomly generated – thus, clusters of the same color in different graphs do not necessarily correspond to one another.

**Supplementary Figure 4.**
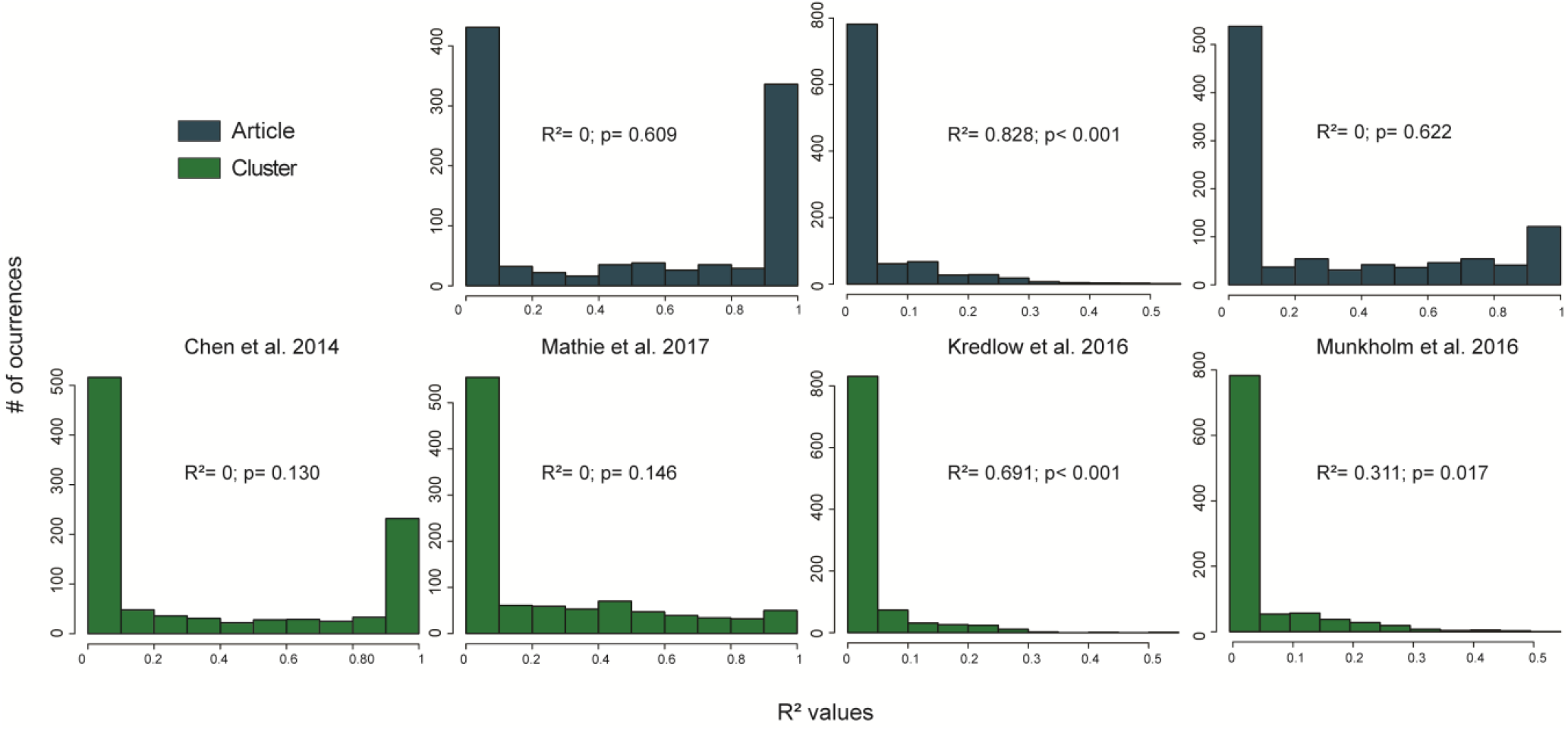
R^2^ probability density distributions. For each meta-analysis, we randomly shuffled results among subgroups, while maintaining the subgroup structure for each meta-analysis in terms of subgroup number and size. For each reshuffling, we calculated R^2^ using a mixed-effects meta-regression model. Histograms show the random R^2^ probability distributions when grouping by article or authorship cluster using the structure of each meta-analysis, which were used to estimate the p-value for the actual R^2^ values displayed in Table 2. For Chen et al. R^2^ calculation for article grouping was not necessary, as each article contributed a single result.

**Supplementary Figure 5.**
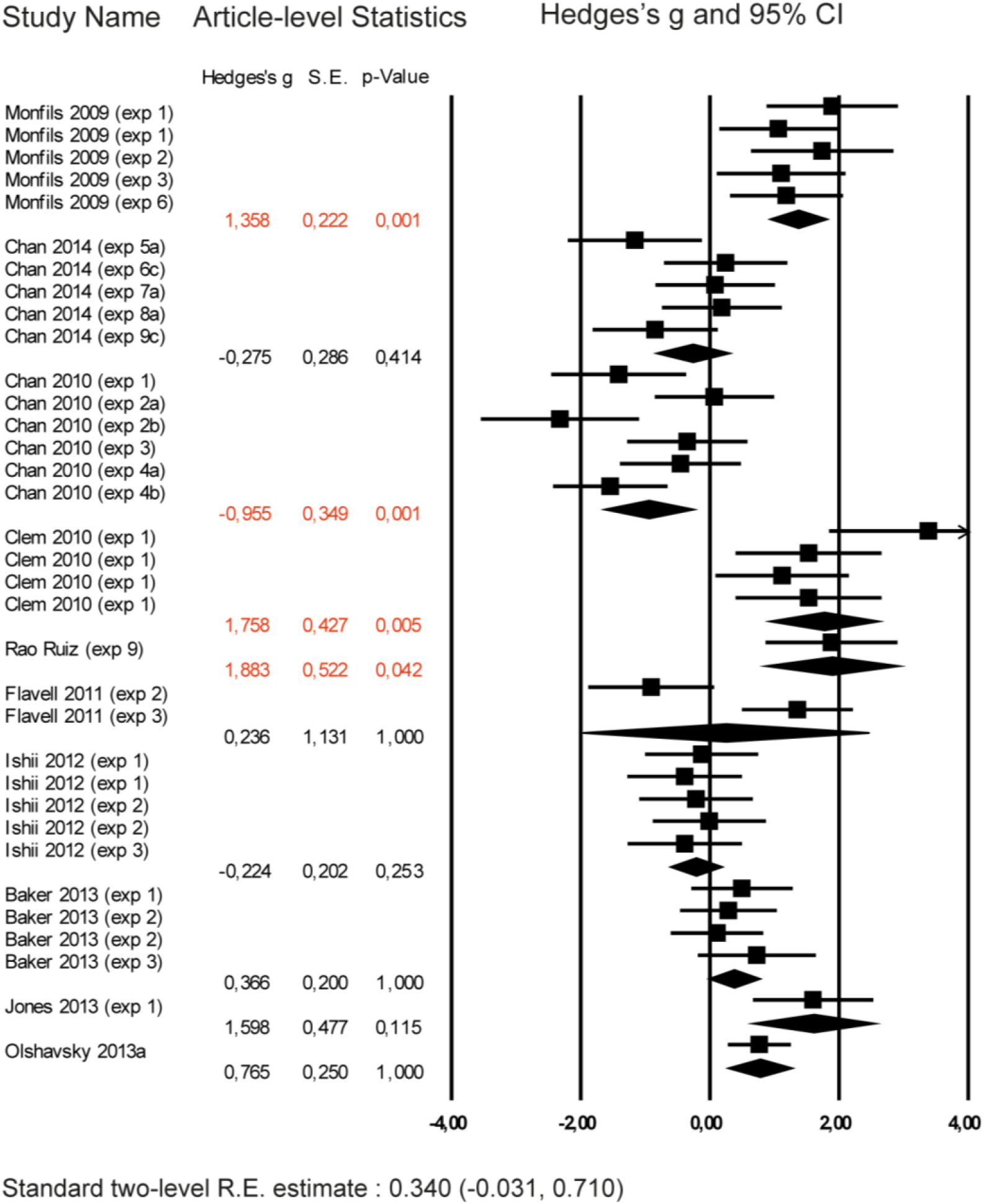
Outcome effect sizes from Kredlow et al. grouped by article. As the meta-analysis from Kredlow et al. was the only one to indicate influence of article-level grouping on heterogeneity, individual outcome effect sizes were grouped for visualization of trends within studies. The plot shows the effect size (in Hedges’ g) and error (95% CI) of individual outcomes (squares), andthe estimates of meta-analyses for each article (diamonds). Each subgroup was compared against the remaining studies within the meta-analysis by a Wald-type test, yielding Bonferroni-corrected p-values shown on the right column. Estimates and corrected p-values of clusters significantly differing from the rest of the meta-analysis at an α of 0.05 are shown in red.

## Notes

### Competing Interest Statement

The authors have declared no competing interest.

### Funding Statement

Fundacao Carlos Chagas Filho de Amparo a Pesquisa do Estado do Rio de Janeiro (FAPERJ), Grant/Award Numbers: E-26/201.544/2014 and E-26/203.222/2017; and by scholarships of the Conselho Nacional de Desenvolvimento Cientifico e Tecnologico (CNPq)

### Author Declarations

All relevant ethical guidelines have been followed and any necessary IRB and/or ethics committee approvals have been obtained.

Any clinical trials involved have been registered with an ICMJE-approved registry such as ClinicalTrials.gov and the trial ID is included in the manuscript.

